# Comparative Cohort Study of Post-Acute Covid-19 Infection with a Nested, Randomized Controlled Trial of Ivabradine for Those With Postural Orthostatic Tachycardia Syndrome (The COVIVA Study)

**DOI:** 10.1101/2023.04.25.23289110

**Authors:** David Saunders, Thomas B. Arnold, Jason M. Lavender, Daoqin Bi, Karl Alcover, Lydia D. Hellwig, Sahar T Leazer, Roshila Mohammed, Bethelhem Markos, Kanchana Perera, Dutchabong Shaw, Priscilla Kobi, Martin Evans, Autumn Mains, Marian Tanofsky-Kraff, Emilie Goguet, Edward Mitre, Kathleen P Pratt, Clifton L Dalgard, Mark C Haigney

## Abstract

**Background:** Significant clinical similarities have been observed between the recently Described ‘Long-Haul’ COVID-19 (LHC) syndrome, Postural Orthostatic Tachycardia Syndrome (POTS) and Inappropriate Sinus Tachycardia (IST). Shared symptoms include light-headedness, palpitations, tremulousness, generalized weakness, blurred vision, chest pain, dyspnea, “brain-fog”, and fatigue. Ivabradine is a selective sinoatrial node blocker FDA-approved for management of tachycardia associated with stable angina and heart failure not fully managed by beta blockers. In our study we aim to identify risk factors underlying LHC, as well as the effectiveness of ivabradine in controlling heart rate dysregulations and POTS/IST related symptoms.

**Methods/Design:** A detailed prospective phenotypic evaluation combined with multi-omic analysis of 200 LHC volunteers will be conducted to identify risk factors for autonomic dysfunction. A comparator group of 50 volunteers with documented COVID-19 but without LHC will be enrolled to better understand the risk factors for LHC and autonomic dysfunction. Those in the cohort who meet diagnostic criteria for POTS or IST will be included in a nested prospective, randomized, placebo-controlled trial to assess the impact of ivabradine on symptoms and heart rate, assessed non-invasively based on physiologic response and ambulatory electrocardiogram. Additionally, studies on catecholamine production, mast cell and basophil degranulation, inflammatory biomarkers, and indicators of metabolic dysfunction will be measured to potentially provide molecular classification and mechanistic insights.

**Discussion:** Optimal therapies for dysautonomia, particularly associated with LHC, have yet to be defined. In the present study, ivabradine, one of numerous proposed interventions, will be systematically evaluated for therapeutic potential in LHC-associated POTS and IST. Additionally, this study will further refine the characteristics of the LHC-associated POTS/IST phenotype, genotype and transcriptional profile, including immunologic and multi-omic analysis of persistent immune activation and dysregulation. The study will also explore and identify potential endotheliopathy and abnormalities of the clotting cascade.

**Trial registration:** ClinicalTrials.gov, ID:NCT05481177 Registered on 29 July 2022.

## Background

### Introduction & Significance

Between 13-70% of coronavirus disease 2019 (COVID-19) survivors continue to experience symptoms after >12 weeks of resolution of primary illness. This phenomenon has been termed “Long Haul COVID-19” (LHC). There is a plethora of reported LHC symptoms including dizziness, tachycardia, palpitations, confusion, fatigue, memory loss, dyspnea and many more (1).^i^ LHC symptoms greatly degrade an individual’s quality of life and functional ability. The Postural Orthostatic Tachycardia Syndrome (POTS) sometimes referred to as Soldier’s Heart or DaCosta’s Syndrome is a particularly debilitating condition (2). POTS overlaps with several of the clinical features of LHC, particularly tachycardia, confusion, and fatigue. Prior to COVID- 19, POTS was a relatively rare phenomenon, affecting otherwise healthy individuals following viral illness or trauma. Recent reports suggest that a substantial number of LHC patients meet diagnostic criteria for POTS (3). Clinics dedicated to the treatment of autonomic disorders are seeing substantially increased numbers of POTS patients among those who have recovered from COVID-19. The American Autonomic Society issued a statement alerting clinicians to consider the possibility of POTS when assessing LHC patients (4). However, the actual prevalence of those with LHC also meeting the diagnostic criteria for POTS has been only minimally characterized. Inappropriate Sinus Tachycardia (IST) is a related syndrome associated with sinus heart rates that are elevated above 100 beats per minute at rest and an average rate >90 over a 24- hour period. The symptoms associated with IST are similar to those of POTS.

The symptoms of LHC have been described in prospective studies. In a prospective study of 312 individuals diagnosed with COVID during the first pandemic wave in Norway, 55% reported symptoms 6 months following primary infection. The most common symptoms were fatigue (30%), disturbed taste and/or smell (27%), concentration (19%) or memory (18%) problems, dyspnea (15%), headache (11%), and palpitations (6%) (5). These symptoms are similar to those reported by POTS sufferers, which include light-headedness, palpitations, tremulousness, generalized weakness, blurred vision, chest pain, dyspnea, “brain-fog”, and fatigue (6). Limited systematic data exist regarding the hemodynamic, autonomic, or metabolic phenotypes in LHC, or corresponding genotypic risk factors.

Consensus diagnostic criteria for POTS has been endorsed by numerous professional societies (7). These criteria include chronic symptoms and a defined sustained increase in heart rate after standing or head-up tilt. This heart rate increase should not be attributable to orthostatic hypotension or other conditions that may provoke sinus tachycardia. Additionally, the demonstrated increase in heart rate must coincide with the symptoms in question (such as light headedness, palpitations, tremulousness, generalized weakness, blurred vision, and fatigue, vasovagal syncope). Additionally, consensus statements define diagnostic criteria for IST as the presence of both average and absolute elevated resting sinus heart rate (8).

The present study will aim to further define risk factors for LHC and POTS/IST, better understand the underlying pathophysiology, estimate the proportion of individuals presenting with POTS symptoms who have the diagnosis, and evaluate an intervention (ivabradine) in a nested, randomized trial in those found to have POTS.

### Evidence for LHC/POTS and Immunologic Dysregulation

While increasingly common, the pathophysiological mechanisms driving post-COVID-19 cardiovascular and autonomic complications, including tachycardia, remain unknown (9). Nalbandian *et al* suggest that possible mechanistic pathways responsible for LHC include 1) residual damage of cells and tissues caused directly by the SARS-CoV-2 infection, 2) expected long-term damage due to critical illness, and 3) persistent immunological aberrations (10). Several mechanisms have been postulated by which immune dysfunction may contribute to LHC. First, sustained inflammation, triggered by persistent virus, viral antigens, or immune dysregulation triggered by the initial infection, could directly cause cardiac damage (11). Persistent immune activation can lead to cardiac fibrosis, and prolonged exposure to pro inflammatory cytokines can prolong ventricular action potentials (12). Significantly for this study, chronic inflammation can also affect autonomic pathways (10). Second, individuals with LHC may develop T-cell and/or natural killer (NK) cell exhaustion, defined as reduced cellular responses to SARS-CoV-2, and this can then lead to viral persistence and the symptoms of LHC. Finally, COVID-19 may trigger development of autoantibodies, including antiphospholipid antibodies or antibodies against adrenergic and muscarinic receptors, that could then contribute to the cardiovascular manifestations of LHC (11, 13). Immunologic mechanisms discussed in the literature include persistent immune activation, T-cell/NK cell exhaustion, and development of autoantibodies (10-14). The study will investigate whether other immune mechanisms play a role in contributing to post-COVID-19 POTS. Alterations in basophil and mast cell responsiveness may influence development of POTS syndrome. One of the possible mechanisms underlying POTS is activation of basophils and mast cells by parasympathetic neurons. This hypothesis is supported by a recent study where two thirds of individuals with POTS demonstrated increased histamine or other mast cell mediators in the blood in the setting of postural tachycardia (15). The present study will investigate whether this also occurs in post COVID-19 POTS. Biomarkers of mast cell and basophil degranulation, including histamine, tryptase, and prostaglandin D2, will be measured before and after orthostasis testing. Additionally, participants will be evaluated for increased basophil sensitivity to activation by calcitonin gene-related peptide (CGRP) and substance P at baseline - molecules released by parasympathetic nerves that can stimulate mast cell and basophil activation. Immune complexes may be important drivers of persistent inflammation after resolution of SARS-CoV-2 infection. Immune complexes are known to develop in the context of SARS-CoV-2 infection, and it has been demonstrated they are able to persist in tissues and drive inflammation despite the resolution of viral infection (16, 17). Such immune complexes can interact with the mast cell via Fcγ-receptor. These complexes interact with a host of other immunological systems and affect NK cell activity. NK cell responsiveness to immune complexes may be measured as well if indicated.

### Evidence for LHC/POTS and Hematologic Dysfunction

In addition to immunologic dysregulation and cytokine storm, blood coagulation and anticoagulant pathways, as well as fibrinolysis, are perturbed in COVID-19, increasing risks of both macro/ micro-thrombosis and hemorrhagic pathologies with increasing infection severity (18). Plasma or serum levels of fibrin D-dimer, a product of fibrinolysis, are now routinely measured in clinical assays for COVID-19 patients, while assays for additional up or down regulated pro and anti-coagulant proteins and their activities are still largely restricted to research laboratories. For example, the acute-phase protein von Willebrand factor (VWF) is highly up-regulated in severe COVID-19, concomitant with reduced activity of the protease ADAMTS13, which cleaves VWF following its secretion from endothelial cells or platelets, thereby reducing its prothrombotic potential (19, 20). Persistent endothelial damage is a feature of LHC (21). Emerging evidence indicates that endotheliopathy and a prothrombotic phenotype also persist in LHC (22). This includes altered thrombin lag times, endogenous potential and peak levels, as well as up-regulated VWF, factor VIII (FVIII) and thrombomodulin (23).

A recently published preprint reported that gamma prime fibrinogen (GPF), which is translated from a splice variant of fibrinogen, is highly up-regulated in severe COVID-19 (22). Interestingly, GPF binds to the heparin-binding exosite on thrombin, preventing its neutralization by heparin and antithrombin III and increasing resistance of clots to fibrinolysis (24). It is hypothesized that GPF, like VWF and FVIII, may also be up-regulated in LHC. Further characterizing altered hemostatic profiles in LHC could suggest more effective measures to prevent thrombosis, e.g., potential effectiveness of direct thrombin inhibitors in patients with elevated GPF and/or who have failed low-molecular weight heparin therapy.

### Psychosocial, Quality of Life, and Cognitive Factors

Despite a growing number of studies documenting the clinical impact of LHC, few have directly examined the broad psychosocial and cognitive functioning impairments that may affect individuals who suffer from the condition (25). COVID-19, in the absence of the long-haul syndrome, has been associated with numerous psychological symptoms including, but not limited to psychological distress, post-traumatic stress disorder, secondary traumatic stress, complicated grief, and anxiety (26-29). Not surprisingly, symptoms reported in the association with LHC are remarkably similar to many of the criteria for mood and anxiety disorders described in the Diagnostic and Statistical Manual of the DSM, 5th edition (DSM-5) (30). For example, heart palpitations, cognitive impairment, headache, sleep disturbance, dizziness, delirium, reduced appetite, and fatigue are also often present in several DSM-5 disorders. To date, preliminary research on adults with LHC suggests that depression/anxiety, cognitive impairment, loss of concentration, fatigue, and insomnia are common (31, 32). Importantly, this research has also found that participants reported a significantly diminished quality of life. While there are clearly physiological underpinnings of COVID-19 contributing to symptoms during acute illness, given the links between psychological functioning and COVID-19 that resolves, the extent to which LHC is related to psychosocial and cognitive functioning warrants further exploration.

### Genetic and Phenotypic Assessments

Preliminary work has elucidated genetic mechanisms responsible for COVID-19 pathophysiology. The involvement of the cell surface receptor angiotensin converting enzyme 2 (ACE2) and the transmembrane protease serine-2 (TMPRSS2) protein in binding and cleavage of SARS-CoV-2 has been reported in multiple studies (33). The association between the polymorphisms of renin-angiotensin-aldosterone system-related genes, i.e., ACE2 and TMPRSS2, and the severity of COVID-19 disease have been thoroughly investigated (34). Studies have correlated single nucleotide polymorphisms (SNPs) of ACE2 and TMPRSS2 with lower severity of COVID-19 in an Indian population (35, 36), and a lower risk of developing severe COVID-19 (37). Another meta-analysis found that a TMPRSS2 SNP was not associated with more severe COVID-19 manifestations, whereas two ACE2 SNPs were associated with more severe COVID-19 manifestations by genotype but not allele (38).

A recent study identified that the ACE2 T allele was associated with a higher risk for critical COVID-19 o ACE2 with a higher risk of severe COVID-19 (41). Given ACE2 and TMPRSS2 polymorphism associations with severe COVID-19, the study will investigate associations with post-COVID symptoms including fatigue and dyspnea. These polymorphisms influence variability of ACE2 and TMPRSS2 expression with the ACE2 rs2285666 polymorphism elevating ACE2 expression up to 50%. Higher expression of ACE2 can trigger vascular constriction, worsening endothelial dysfunction, fibrosis and inflammation (42, 43).

Reports have noted elevated plasma ACE2 activity three months after the acute infection in individuals with long-COVID as compared to uninfected matched controls (44). Long-lasting systematic inflammation after acute SARS-CoV-2 is associated with more severe post-COVID symptoms (45). These associations could underlie post-COVID symptoms (44). It is plausible that intron/missense variants of ACE2 and TMPRSS2 polymorphisms could be associated with LHC development.

Most studies investigating the role ACE2 or TMPRSS2 SNPs have focused on the risk of being infected or the severity of COVID-19 disease in the acute phase. Two recent studies investigated the association between five COVID-19 associated SNPs in AC1, ACE2 and TMPRSS2 and long term post-COVID symptoms in previously hospitalized COVID-19 survivors (46, 47). However, the SNPs assessed, although previously associated with COVID-19 severity, did not predispose for developing LHC symptoms in people who were previously hospitalized due to COVID-19. To date, data on genomic associations and predispositions with the development of LHC remain limited. Strong correlation between disease severity and predisposition to LHC have not been established. Genetic susceptibility to endophenotypes overlapping with LHC may contribute to the condition spectrum. The study aims to evaluate polygenic risk factors across medical phenotypes, including cardiovascular, hematologic, immunologic and neurological conditions, for enrichment in POTS/IST subjects from whole genome sequencing and multi-omic signature analysis. Multi-omic profiling aims to enable molecular classification of POTS/IST subgroups from transcriptome and proteome expression of key host and disease factors.

### Management of POTS

Postural Orthostatic Tachycardia Syndrome (POTS) occurs frequently in patients with LHC. There is currently no agreed upon standard of care for POTS. Rate control medications have been studied, as have exercise training and other medications. Low-dose propranolol was previously recommended based on a trial performed at Vanderbilt. A subsequent study by a different group, however, found no improvement in quality of life when using propranolol (48). Propranolol, the recommended agent, is associated with penetration of the central nervous system resulting in nightmares and depression. Bronchospasm is a common (and sometimes life threatening) complication.

In a recent review of POTS (2020), the Canadian Cardiovascular Society acknowledged that case series of ivabradine in POTS have been reported to have 68-78% effectiveness with respect to controlling heart rate and symptoms (49). Ivabradine is a selective sinoatrial node blocker FDA-approved for management of tachycardia associated with stable angina and heart failure not fully managed by beta blockers. It blocks the hyperpolarization-activated cyclic nucleotide-gated (HCN) channel responsible for the cardiac pacemaker I_funny_ current, which regulates heart rate. In clinical electrophysiology studies, cardiac effects were most pronounced at the sinoatrial (SA) node, with lesser prolongation of the AH and PR intervals. There was no effect on ventricular repolarization or myocardial contractility. Ivabradine causes a dose-dependent reduction in heart rate in proportion to the baseline heart rate. Ivabradine’s relative lack of effect on the respiratory system may avoid exacerbating dyspnea experienced by LHC patients. This has been confirmed in a small RCT of patients with postural orthostatic tachycardia syndrome (POTS) where it was shown to safely reduce heart rate (50). Other treatments, including salt supplementation, midodrine, and fludricortisone may soon be superseded by this novel therapy. IST is a similarly intractable clinical condition that is believed to be responsive to ivabradine, although the drug does not have FDA approval for that indication.

## Methods/Design

### Aims and Objectives

This prospective study of 200 LHC volunteers with a nested RCT will assess the effect of ivabradine on heart rate and symptoms in those meeting diagnostic criteria for POTS or IST. A comparator group of 50 volunteers with documented COVID-19 but without LHC will help identify the risk factors for LHC and autonomic dysfunction. The comparator group will also be evaluated for POTS and eligible for inclusion in the nested RCT.

The primary aim is to identify the proportion of volunteers with relevant symptoms diagnosed with postural orthostatic tachycardia syndrome (POTS) or inappropriate sinus tachycardia (IST). We will first attempt to address the frequency of clinically confirmed POTS in those with persistent COVID-19 symptoms suggestive of autonomic dysfunction. The second aim is to determine the potential benefits of ivabradine treatment for those with LHC-associated POTS or IST. A nested clinical trial for Ivabradine responsiveness on reduction of tachycardia. Ivabradine is a drug approved to treat tachycardia in severe heart failure with some demonstrated improvement in POTS outcomes.^ii^ Those with confirmed POTS or IST will be randomized to ivabradine or placebo to determine efficacy in reducing heart rate and POTS symptoms.

The third aim is to characterize potential differences in hematologic, metabolic, immunologic, and genetic variants among volunteers with and without LHC. Cellular and molecular characterization of LHC and non-LHC participants will be performed. This aim will identify risk factors and provide mechanistic insights into the pathophysiology of LHC and POTS for those with and without LHC. The biopsychosocial mechanisms of LHC and POTS will also be explored. Study objectives are summarized in Table 1.

**Table 1.** **COVIVA Study Objectives.**

| Objectives | Study Endpoints/Milestones |
| --- | --- |
| 1. Assess frequency and severity of fatigue, dyspnea, headache, confusion and other suggestive symptoms in volunteers with and without LHC; measure occurrence of clinically confirmed POTS or IST—in relation to symptoms | Proportions of LHC and non-LHC volunteers with POTS; disease severity based on composite physiologic (ambulatory EKG, vital signs, orthostatics, Objective 3) and well-being assessments (Objective 5) |
| 2. Assess whether treatment with ivabradine will significantly reduce heart rate; improve symptoms and quality of life among volunteer with POTS or similar dysautonomia | Primary endpoint for the nested ivabradine RCT - reduction in standing heart rate at three months. A small cross-over RCT in POTS patients without COVID-19 history found ivabradine reduced standing heart rate ~16 bpm between treated and placebo groups (see text) |
| 3. Determine whether wearable automated ambulatory monitoring with existing FDA-approved devices will improve diagnostic accuracy for POTS | The differential proportion of individuals diagnosed with POTS using conventional (in-office) and wearable diagnostic methods |
| 4. Determine the degree to which LHC induces autonomic instability and its associations with induced inflammation and/or dysregulation | Degree and severity of POTS as in Objectives 1 and 3. Levels of circulating catecholamines before and after orthostatic maneuvers |
| 5. Evaluate potential contribution of psychological symptoms and interplay between psychosocial and cognitive symptoms, quality of life, and physiologic responses induced by LHC | Changes in quality of life (SF-36); psychosocial functioning (PHQ-SADS, DASS-21 surveys); Structured Clinical Interview for DSM-5 disorders; cognitive function (PROMIS, NIH Toolbox) |
| 6. Determine if LHC improves following subsequent vaccination and/or “breakthrough” COVID-19 infection | Changes in quality of life, symptom surveys and/or laboratory markers of infection in those vaccinated or contracting infection during the study |
| 7. Compare antibody and/or cellular immune responses to SARS-CoV-2 antigens in LHC patients and infected individuals without LHC | 1. IgG antibodies to SARS-CoV-2 or the four major seasonal human coronaviruses (HCoV-OC43, HCoV-HKU1, HCoV-229E, or HCoV-NL63)<br>2. T-cell cytokine responses against SARS-CoV-2<br>3. Evaluate whether individuals with LHC have decreased immunoregulatory networks compared to infected individuals without LHC<br>4. Assess NK cell phenotypic/functional differences<br>5. Identify if POTS-related orthostasis is associated with mast cell degranulation and basophil activation product release (histamine, tryptase)<br>6. Evaluate whether basophil responsiveness to activation in response to mediators released by parasympathetic nerves (Calcitonin gene-related peptide (CGRP), Substance P) is different between individuals with POTS and those without. |
| 8. Assess whether markers of dysregulated blood coagulation (significantly correlated with immunothrombosis in severe COVID-19) persist in individuals with LHC | PT, APTT, factor VIII:C, total fibrinogen, D-dimer, thrombin generation, thromboelastometry (ROTEM); Exploratory - gamma prime fibrinogen, von Willebrand factor (VWF) antigen, VWF propeptide, ADAMTS13 activity, soluble thrombomodulin |

**Table 1. COVIVA Study Objectives.**
| # | Objective | Study Endpoints/Milestones |
| --- | --- | --- |
| 1. | To assess the frequency and severity of fatigue, dyspnea, headache, and confusion and other suggestive symptoms among volunteers with and without LHC and measure the occurrence of clinically confirmed POTS or IST—in relation to these symptoms | Proportions of LHC and non-LHC volunteers with postural orthostatic tachycardia syndrome (POTS), and disease severity based on a composite of physiologic (ambulatory EKG, vital signs, orthostatics, see Objective 3) and well-being assessments (see Objective 5). |
| 2. | To assess whether treatment with ivabradine will significantly reduce heart rate and improve symptoms and quality of life among subjects manifesting typical POTS or similar autonomic dysregulation | The primary endpoint of the nested ivabradine RCT will be a reduction in standing heart rate at three months. Based on a prior small cross-over RCT in POTS patients without a history of COVID-19, a reduction of roughly 16 bpm in standing heart rate between the treated and placebo groups was observed.<sup>1</sup> |
| 3. | To determine whether wearable automated ambulatory monitoring with existing FDA-approved devices will permit improved diagnostic accuracy for POTS. | The differential proportion of individuals diagnosed with POTS using conventional (in-office) and wearable diagnostic methods. |
| 4. | To determine the degree to which LHC induces autonomic instability and whether it is associated with induced inflammation and/or dysregulation | Degree and severity of POTS as in Objectives 1 and 3. Levels of circulating catecholamines measured in the blood before and after orthostatic maneuvers. |
| 5. | To evaluate potential contribution of psychological symptoms and the interplay between psychosocial symptoms, quality of life, cognitive symptoms, and physiologic responses induced by LHC. | Improvements/changes in quality of life measured by the SF-36, psychosocial functioning measured by the PHQ-SADS and DASS-21 surveys, and cognitive function measured by the PROMIS cognition survey and NIH Toolbox tasks. |
| 6. | To determine whether LHC improves following vaccination (in the limited number of cases where late vaccination occurs) and/or “breakthrough” clinical COVID-19 infection. | Changes in longitudinal quality of life and symptoms surveys and/or laboratory markers of infection in those receiving vaccination or contracting infection during the study. |
| 7. | Assess whether individuals with LHC have different antibody and/or cellular immune responses to SARS-CoV-2 antigens compared to infected individuals who did not develop LHC | 1. Assess whether IgG antibodies to SARS-CoV-2 or the four major seasonal human coronaviruses (HCoV- OC43, HCoV-HKU1, HCoV-229E, or HCoV-NL63) are associated with LHC.<br>2. Assess if T-cell cytokine responses against SARS- CoV-2 proteins are associated with LHC.<br>3. Evaluate whether individuals with LHC have decreased immunoregulatory networks compared to infected individuals who did not develop LHC.<br>4. Assess NK cell phenotypic/functional differences in LHC and non-LHC volunteers.<br>5. Identify if orthostasis in POTS disease is associated with release of mast cell degranulation and basophil activation products such as histamine and tryptase, and evaluate whether basophil responsiveness to activation in response to mediators released by parasympathetic nerves such as Calcitonin gene-related peptide (CGRP) and Substance P is different between individuals with |
|  |  | POTS disease and those without. |
| 8. | Assess whether specific plasma protein markers of dysregulated blood coagulation, which were significantly correlated with immunothrombosis in severe COVID-19, persist in individuals with LHC | Clinical or research-grade laboratory assays will quantify and characterize specific markers of dysregulated coagulation and immunothrombosis. Assays will include PT, APTT, factor VIII:C, total fibrinogen, D-dimer, thrombin generation assay and thromboelastometry (ROTEM) assay. Additional analytes may include gamma prime fibrinogen, von Willebrand factor (VWF) antigen, VWF propeptide, ADAMTS13 activity and soluble thrombomodulin. |

### Ethical Approval

The study was approved by the Uniformed Services University of the Health Sciences Institutional Review Board in June 2022 and registered with clinicaltrials.gov as NCT05481177. The study will adhere to the principles of Good Clinical Practice.

### Study design

The study design is shown in Figure 1. The study will enroll a cohort of 200 volunteers with LHC from both the Military Health System (MHS) and civilian populations in the Washington metropolitan area (which includes the District of Columbia and parts of Maryland, Virginia, and West Virginia). In addition, a small comparator group of 50 volunteers with a confirmed COVID-19 diagnosis but without ongoing sequelae will be enrolled in the overall cohort.

**Figure 1.**
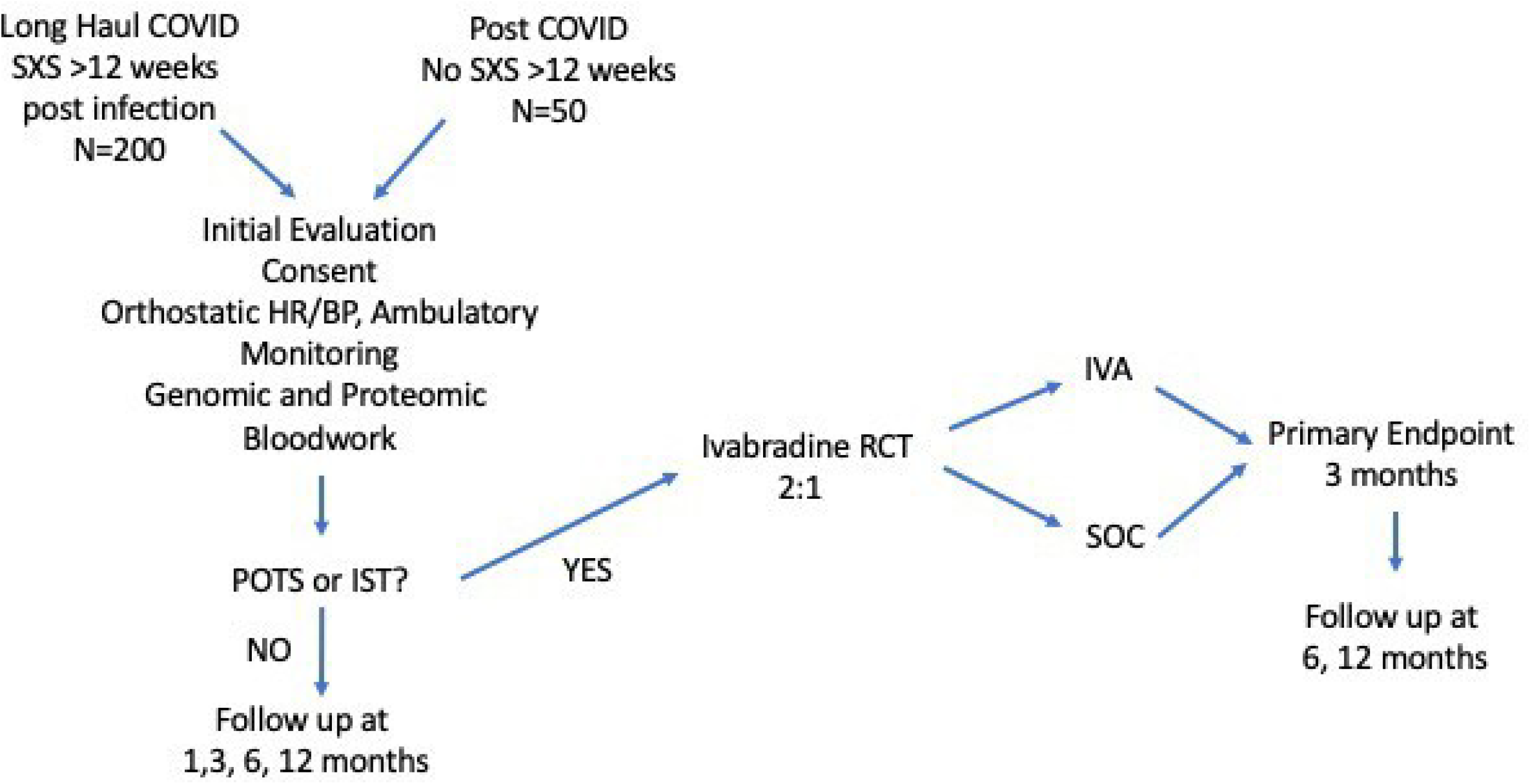
Overall Study Schema. A diagram of the general study design. The study will include an observational cohort with 250 volunteers including 200 with long-haul COVID and 50 controls without LHC. Those from the observational cohort who are diagnosed with POTS will be randomized to a nested, placebo-controlled trial of ivabradine to control heart rate and improve symptoms. (POTS - Postural Orthostatic Tachycardia Syndrome; IST - Inappropriate Sinus Tachycardia; IVA - Ivabradine; SXS - Symptoms; HR - Heart Rate; BP - Blood Pressure).

### Study Site and Population

The study will take place at a single center in suburban Maryland. The cohort will be enriched for volunteers with one or more signs or symptoms consistent with autonomic dysfunction indicating potential POTS/IST during the recruiting process. Examples include dizziness, heart palpitations, fainting spells or racing heart rate. Other non-specific symptoms associated with POTS have included headache, ‘brain fog’ or confusion, gastrointestinal disturbances, and fatigue. Inclusion criteria are: 18-80 years old; History of documented COVID-19 infection of any severity to include a positive COVID-19 PCR, or antibody test; meet criteria for ‘long-haul’ COVID-19 with persistent symptoms >12 weeks following acute illness for the LHC cohort (or without persistent long-haul COVID-19 symptoms for the non-LHC cohort); able and willing to provide informed consent and participate for the 1-year study duration; Willing to participate in the nested Ivabradine RCT if enrollment criteria are met; and access to healthcare and health insurance.

Exclusion criteria are resting heart rate <60 bpm; atrial fibrillation; supraventricular tachycardia; allergic reaction or known contraindications to study drug; pregnant/lactating females; impaired gastrointestinal drug absorption; acute suicidality identified at screening; taking any of the following unless discontinued in consultation with the volunteer’s healthcare provider and a one week washout period: beta-blockers, calcium-channel blockers, cholinesterase inhibitors (pyridostigmine), vasoconstrictors (midodrine, octreotide, droxidopa, stimulants), sympatholytics (clonidine, methyldopa), blood volume enhancers (fludrocortisone, desmopressin, salt supplementation), or oral ketoconazole.

### Detailed Study Methods

Study methods are described in detail in **Figure 2** and the accompanying Spirit Figure (**Figure 3**). Following informed consent, volunteers will be screened to determine whether they meet the study enrollment criteria. Initial screening will include medical history including concomitant medications, and physical exam, structured clinical interview for DSM-5 psychiatric disorders, COVID-19 antibody tests, EKG, baseline spirometry to assess pulmonary function, and clinical safety laboratory testing to include serum chemistries, complete blood count, and urinalysis. Subjects will have thyroid functions tests (TFTs), highly sensitive C-reactive protein (CRP), cardiac specific troponins and pro-BNP levels (the latter to assess for myocarditis) at screening only. Females will have a urine test for β-hCG at screening, and if positive, serum β-hCG (Figure 2 and 3). The initial screening will be valid for 60 days prior to participation in a nested randomized placebo-controlled trial of ivabradine to reduce heart rate and improve symptoms (nested RCT).

**Figure 2.**
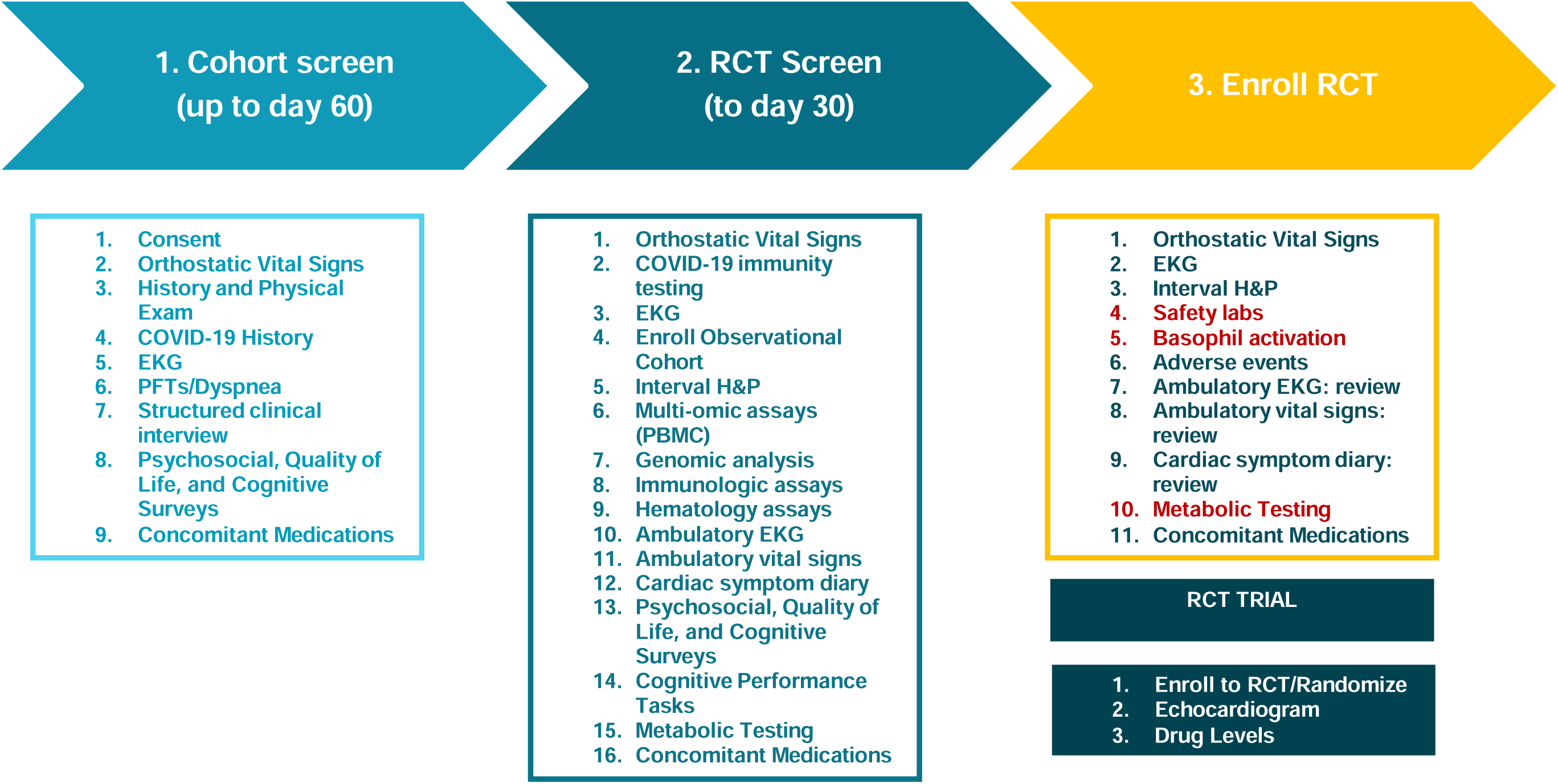
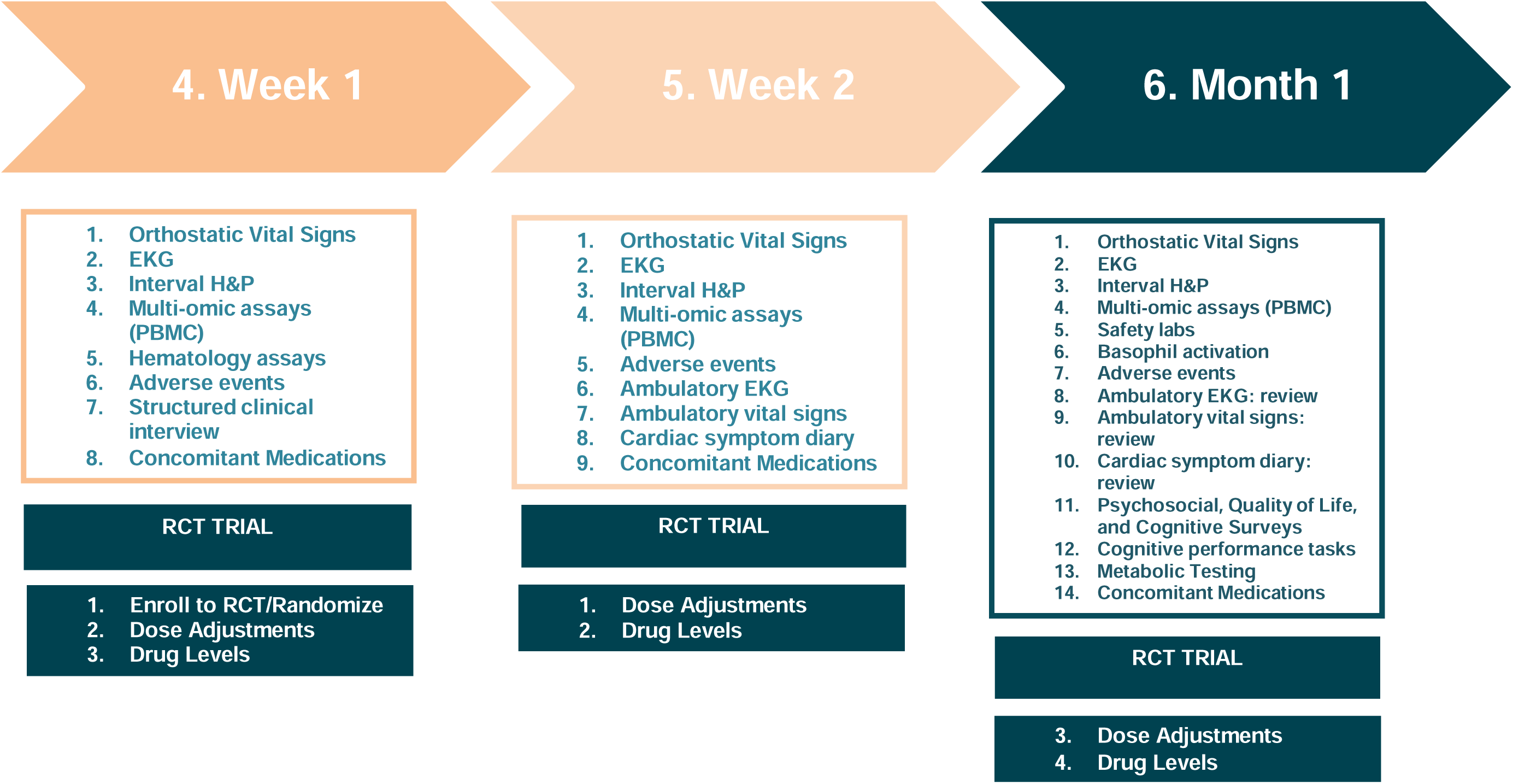
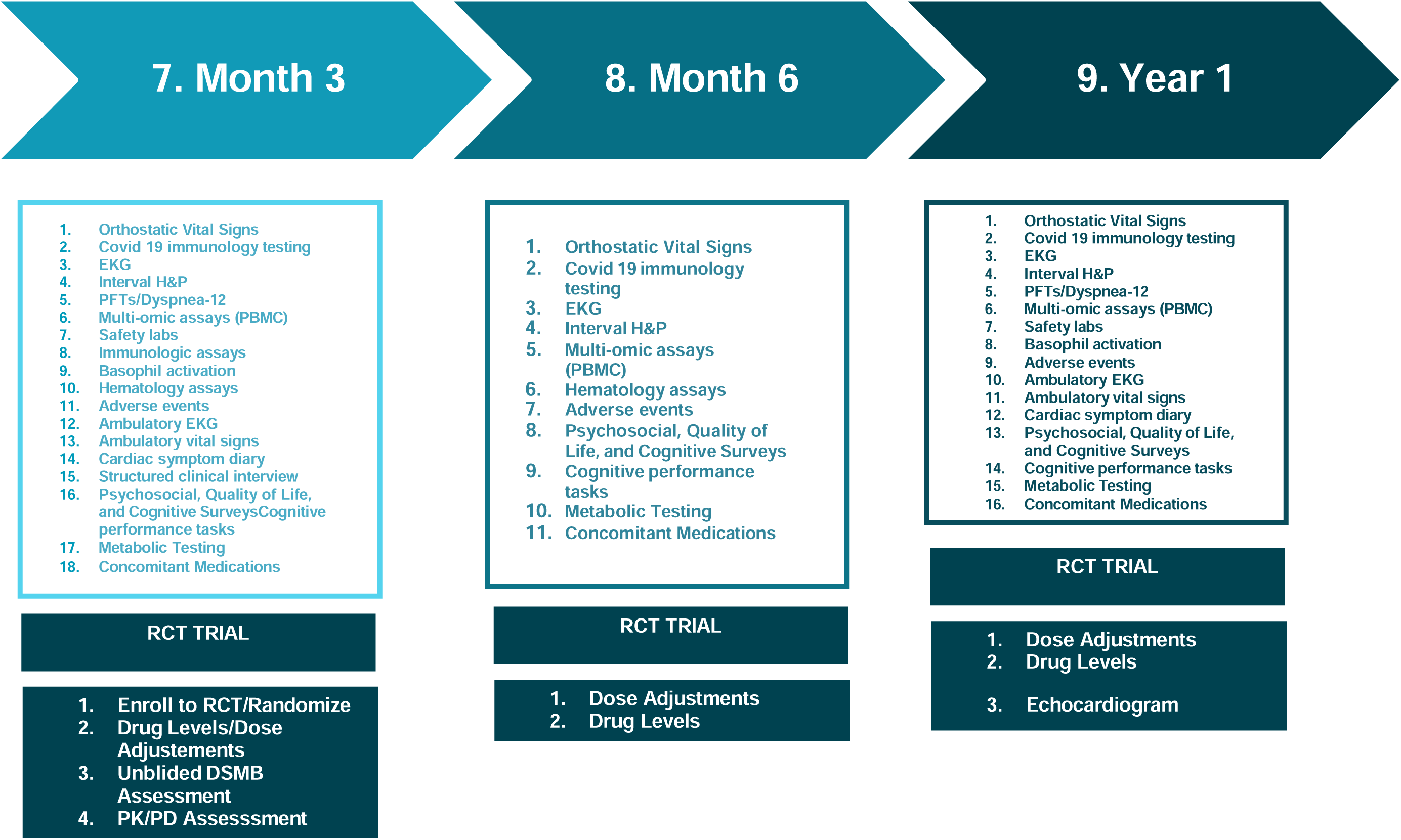
COVIVA Study Schedule of Events. A listing of times and events for study procedures.

**Figure 3.**
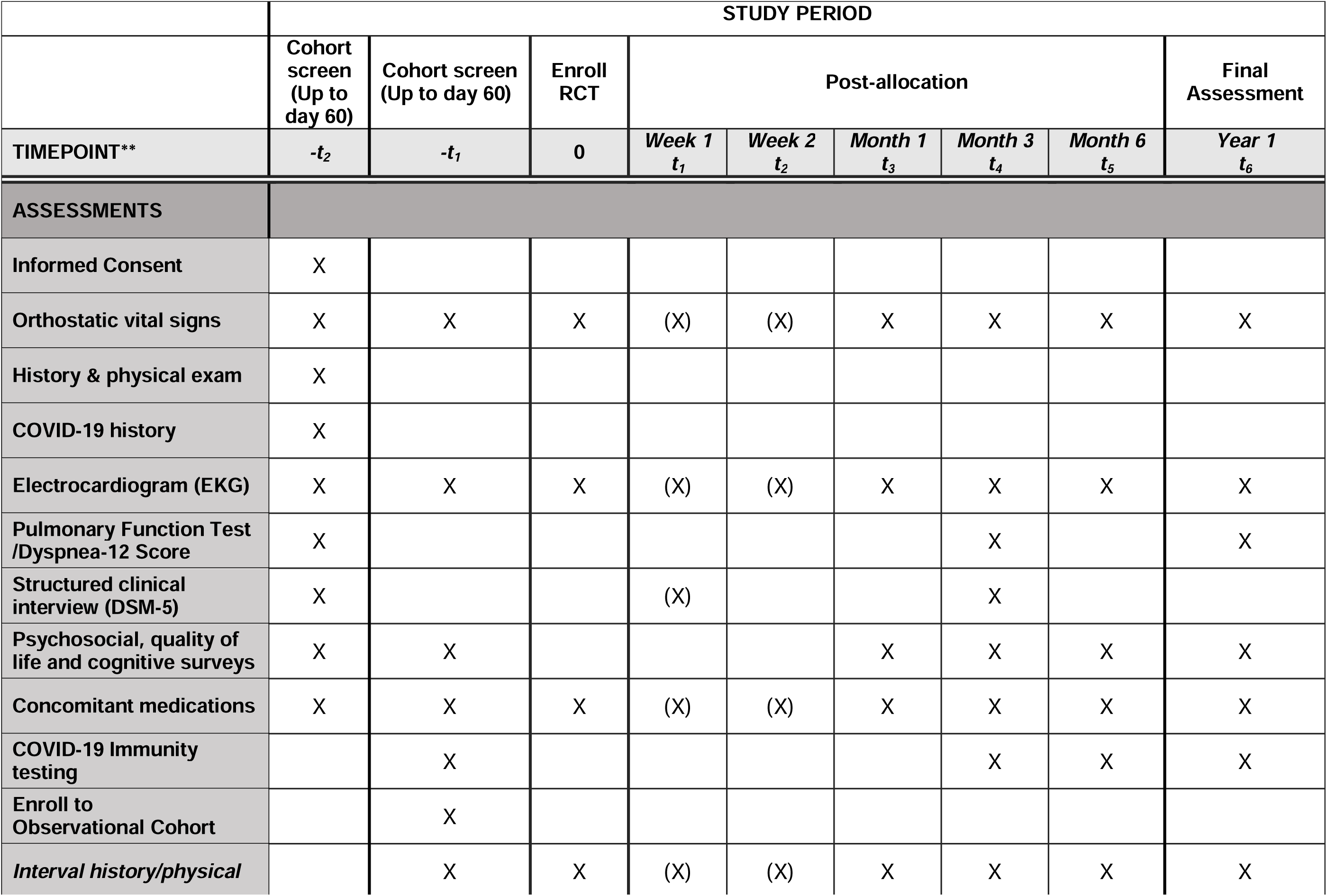
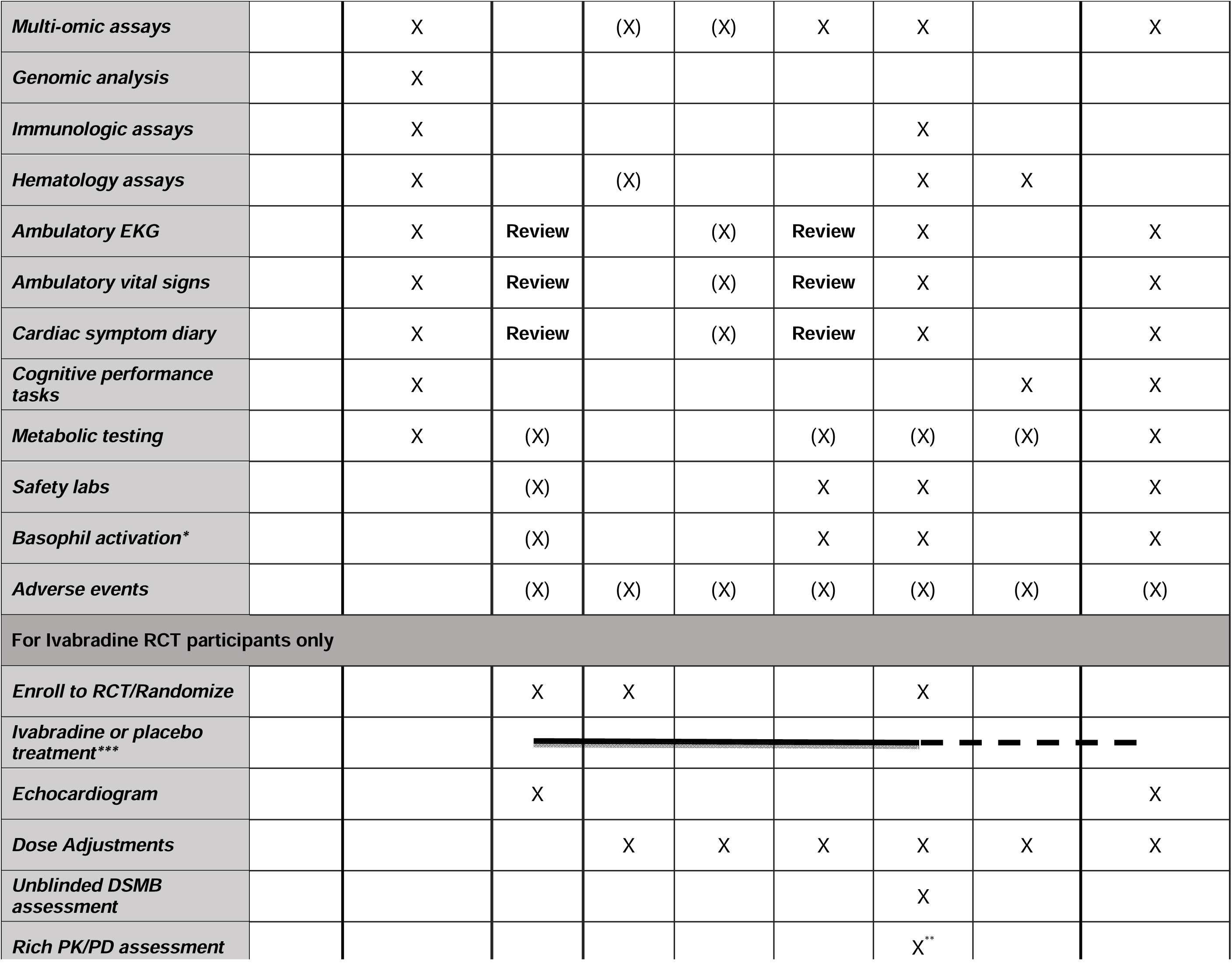
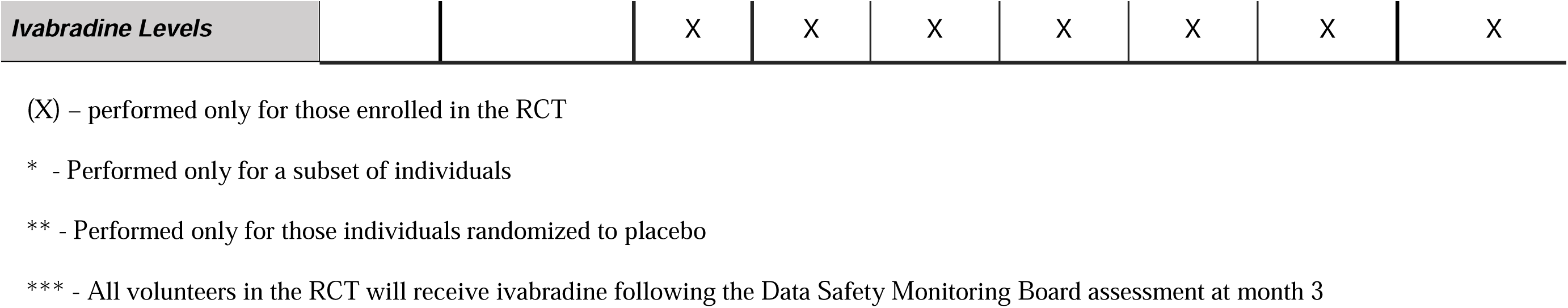
**COVIVA Study Spirit Figure**

After the initial Cohort Screening visit, volunteers meeting criteria will return for enrollment into the overall LHC cohort and screening for the nested RCT (visit 2). Enrolled volunteers will undergo whole genome sequence analysis to look for biomarkers of LHC risk, along with multi omic analysis. Volunteers will have orthostatic vital signs, an interval medical history, EKG, immunologic assays, psychosocial, quality of life, and cognitive surveys; complete cognitive performance tasks; undergo metabolic testing, and have concomitant medications reviewed. Volunteers enrolled to the overall study cohort will be screened for enrollment in the nested RCT for POTS. Volunteers will be provided with ambulatory electrocardiographic and vital signs monitoring devices at enrollment, and complete of a detailed symptom diary for review by study investigators after 1 week of monitoring. Investigators will determine whether volunteers meet criteria to participate in the nested (RCT) of ivabradine to reduce heart rate and improve POTS symptoms. The inclusion criteria for the nested RCT are the same as the overall cohort plus: POTS diagnosed based on increased heart rate from supine to standing ≥ 30 beats per minute and < 20 mm Hg drop in systolic or <10 mmHg drop in diastolic blood pressure. Volunteers diagnosed with IST with or without LHC based on a 24-hour mean heart rate of 90 beats per minute or more (in sinus rhythm) will also be included. Females of childbearing age must be willing to use a highly effective form of contraceptive with <1% failure rate or practice abstinence for the RCT duration.

### Treatment Intervention

Ivabradine (Procoralan) at a starting dose of 5mg twice per day will be compared to placebo in addition to otherwise appropriate medical care. The dose will be adjusted based on individual heart rate response in accordance with the approved product label.

### Randomization, Allocation, and Blinding

Study drug will be allocated to treatment groups by an unblinded research pharmacist. Treatment allocation and randomization will be determined in advance using time-blocked randomization with blocks of three. Allocation will be concealed from the volunteers and study team with treatment assignments predetermined by unblinded personnel not otherwise engaged in study procedures. Test article will be concealed from both subjects and investigators by placing medications at predetermined doses in gel capsules. For active treatment, 2.5mg dose units will be encapsulated with a pharmaceutically inactive compound (e.g. microcrystalline cellulose), while the placebo group will receive capsules with only the inactive compound.

### Data Collection

#### Clinical Assessments

Orthostatic vital signs will be collected with volunteers sitting, standing, and lying down with 5 minutes between each measurement. Head-up tilt-table (HUTT) testing will be performed with the volunteer in the fasted state with heart rate and blood pressure monitoring. HUTT will be considered abnormal if the volunteer has an increase in heart rate of 30 beats per minute over baseline with accompanying symptoms of orthostatic intolerance. A history and physical exam at screening will include demographics (age, sex, race, ethnicity), medical history to include documenting the volunteer’s history of COVID-19, vital signs and clinically significant abnormal physical findings. A brief interval medical history and targeted physical exam to include adverse event assessment will be performed at subsequent visits. Adverse events for those enrolled in the RCT will be assessed for severity and treatment relatedness using the Common Terminology Criteria for Adverse Events (CTCAE) v5.0.

Volunteers will have supine 12 lead electrocardiograms after a five-minute period of rest. Continuous ambulatory blood pressure and electrocardiographic monitoring will be measured using appropriate FDA-cleared device(s). Subjects enrolled to the nested RCT will continue to have these assessments during the study through month 3. Subjects not enrolled to the RCT will have these performed for 1 week starting at the second screening visit for the nested RCT and 1 week at least 2 weeks prior to study discharge. Volunteers will maintain a cardiac symptom diary while they have ambulatory EKG and vital signs monitoring. Spirometry will be performed to assess pulmonary function along with a Dyspnea-12 score which will serve as a key indicator of potential confounding by residual and/or pre-existing respiratory illness. An echocardiogram will be performed at baseline for all those enrolled to the nested RCT to exclude subjects with left ventricular ejection fractions <40%.

#### Dose Adjustment

For those enrolled in the RCT, ivabradine dose will be adjusted based on treatment response according to the label. Those taking placebo will have adjustments made using test article intended to match active treatment based on appearance.

#### Psychosocial, Quality of Life, and Cognitive Factors Assessment

Volunteers will complete a series psychosocial and quality of life of surveys. The Short Form Health Survey (SF-36 v2) includes coherent and easily administered items evaluating quality of life in physical, emotional, and social domains (51). The Patient Health Questionnaire-SADS (PHQ-SADS) is a 30-item self-report measure incorporating items from the PHQ-8 (depression symptoms), GAD-7 (generalized anxiety disorder symptoms), and PHQ-15 (somatic symptoms) (52). Volunteers also will complete the self-report Depression Anxiety Stress Scale-21 (DASS- 21) (53), a questionnaire assessing stress, depression, and anxiety symptoms experienced during the past week. The Patient-Reported Outcomes Measurement Information System (PROMIS) Cognitive Function Short Form-8a will assess perceived cognitive function over the past week (54). Modules from the Structured Clinical Interview for DSM-5 disorders – research version (SCID-5-RV) will be used to evaluate psychiatric disorders (e.g., mood, anxiety, somatic disorders, etc.) and assess possible psychological bases for somatic complaints (55). Finally volunteers will complete the NIH Toolbox Cognition Battery (56), a series of cognitive tasks to assess working memory, episodic memory, immediate recall, processing speed, attention, and executive function. (Figure 2 and 3.)

#### Hematology Assays

Clinical and research laboratory assays (PT, APTT, factor VIII:C, total fibrinogen, D-dimer, thrombin generation assay and thromboelastometry (ROTEM), gamma prime fibrinogen, von Willebrand factor (VWF) antigen, VWF propeptide, ADAMTS13 activity and soluble thrombomodulin) will quantify and characterize specific markers of dysregulated coagulation and immunothrombosis.

#### Genomic Analysis

PAXGene Blood tubes will be collected for genomic and transcriptomic profiling. For whole genome sequencing, genomic DNA will be isolated and used as input for PCR-free library preparation using a ligation-based method on automated liquid handling platforms. Sequencing libraries will be sequenced using Illumina NovaSeq 6000 platforms with paired-end 150bp reads. Raw reads will be demuxed, aligned to a reference genome and genomic variants will be called using a DRAGENv4.0 graph genome and machine learning enabled pipeline. Variants will be annotated and interpreted using a phenotype-award pipeline using Genomiser and used as input for polygenic risk score calculation from selected polygenic score sources from the PGS catalog and used for comparative analysis with control-matched general population scores to identify candidate traits associated with POTS/ITS.

#### Multi-omic Assays

Peripheral blood mononuclear cells (PBMCs) will be collected to assess for transcription, translation and expression of key host and disease factors. Metabolomics will be evaluated along with transcriptome and plasma for proteomics.

#### Immunologic Assays

Serology and cytokines will be assessed in serum, with PBMCs collected for cell-based immunity assays to potentially include studies of T and NK cells. Plasma levels of histamine, tryptase, and prostaglandin D2 will be measured 15 minutes before and shortly after orthostasis testing to assess for mast cell and basophil degranulation. Additionally, biomarker levels including IL-1 receptor antagonist (IL-1Ra), adiponectin, and cholinesterases may be evaluated at baseline while participants are in the supine position to determine if these inflammatory and metabolic markers are altered in individuals with LHC and POTS. These measurements will be made on the same plasma samples but only in the supine position.

#### Basophil Activation in Response to CGRP and Substance P

Basophil Activation in response to CGRP and substance P, as well as basophil sensitivity to activation via the IgE signaling pathway, will be measured before starting the study drug and prior to orthostasis testing. Activation assays may be repeated at the 3-month time point.

#### Pharmacokinetic/Pharmacodynamic (PK/PD) Analysis

Drug levels of ivabradine and relevant metabolites will be analyzed. Results will not be included in the study database until after the first 3 months of the study have been completed for all nested RCT subjects to maintain blinding. For those subjects assigned to placebo, at the completion of 3 months, individuals will be offered cross-over to active drug with participation in a rich-PK sampling study to establish population PK-PD data for ivabradine. The first dose of ivabradine (5mg) will be administered followed by serial blood PK specimens at pre-dose (0 hour; within 1 hour prior to dosing), and at 0.25, 0.5, 1, 2, 4, 8 and 24 hours after dosing.

#### Unblinded Data Safety Monitoring Board (DSMB) Assessment

Individual treatment effectiveness will be assessed on a rolling basis by an independent, unblinded DSMB as nested RCT subjects approach completion of the first 3 months of the study. Placebo-treated volunteers will be offered treatment at month 3 with an additional 9 months of follow-up. Treated subjects will be evaluated for treatment benefit and continued at the identified effective dose as appropriate. The DSMB will include the Research Monitor and 3 physicians.

#### Data Storage and Handling

Study data will be recorded in electronic case report forms (CRFs) using the Research Electronic Data Capture system (REDCap). De-identified data will be directly captured from diagnostic devices including ultrasound images, continuous ambulatory vital signs and/or cardiac telemetry locally stored on password-protected devices.

#### Sample Size Justification and Statistical Analysis

Up to 200 evaluable subjects will be enrolled in the general LHC cohort. Study recruiting will be implemented to enrich for volunteers with common clinical features of POTS. At least 20% of those recruited are expected to have appropriate POTS or IST symptoms for enrollment to the nested RCT (ivabradine vs. placebo). The design is estimated to yield an RCT study population of at least 40 evaluable subjects. Drop-outs in all cohorts may be replaced. With at least 36 subjects completing the RCT at 2:1 allocation of treatment to placebo, the study will have 80% power to identify a difference in standing heart rate at 3 months from 90 bpm in the placebo group compared to 80 bpm in the treated group at the 0.05 significance level.

Data will be analyzed using Stata, R, or another equivalent statistical computing package. Participant demographic and clinical characteristics will be characterized. Percentages and means (with standard deviations) will be calculated for categorical and continuous variables, respectively. We will assess differences in characteristic distribution by group using Chi-square and t-tests (or Fisher’s exact test for small sample sizes) as appropriate. Group differences will be determined for outcomes using multilevel linear (for continuous variables) and logistic regression models. Results will be reported using odds ratios (ORs) for binary outcomes and regression coefficients for continuous outcomes. Association estimates will include 95% CIs. A two-tailed error rate of α = .05 will be the threshold for statistical significance.

## Discussion

Since the initial reports emerged on autonomic dysfunction associated with long-haul COVID- 19, substantial progress has been made in better characterizing associated sequelae. Despite these recent gains, a great deal remains poorly understood regarding how best to evaluate and treat a lengthy list of potential symptoms and syndromes (4, 57). Amidst this quandary, POTS has emerged as the most common form of autonomic dysfunction associated with LHC, with as many as 2.5% of those with LHC affected (58). Pre-COVID, longitudinal studies revealed that while often self-limited, residual POTS symptoms may persist for 5 years or longer.

Despite diagnostic advances, efforts to differentiate POTS from other autonomic syndromes remain imperfect. This is due in part to the wide array of organ systems and functions controlled by the autonomic nervous system (ANS). The most common cardiovascular findings may include tachycardia and hypertension from increased sympathetic tone, orthostatic hypotension from decreased baroreceptor and chemoreceptor reflex sensitivity, and reduced heart rate variability (59). Commonly recommended clinical diagnostic tests to evaluate autonomic dysfunction include Valsalva maneuvers, supine and standing vital signs measurements or head-up tilt table testing (HUTT), a 6-minute walk test, deep breathing, and ambulatory EKG testing. Other tests including skin biopsies for evaluation of peripheral nerve inflammation, and emerging wearable device measurements have also been suggested. A range of questionnaires have also been employed including the COMPASS-31, Vanderbilt Orthostatics Symptom Score (VOSS), PROMIS-29, and others (58). In the present study, we have selected a subset of tests in the interests of time, simplicity, and the ability to enroll and retain subjects. Planned tests include HUTT (or supine/standing vital signs for those unable to complete HUTT), ambulatory EKG monitoring, and the COMPASS-31 survey for autonomic dysfunction. Volunteers will be monitored using an advanced automated ambulatory vital signs monitor recently approved by FDA in order to aid in diagnosis (60).

Difficulties in diagnosis can compound the disability and demoralization suffered by POTS patients. An estimated 10% of POTS sufferers may be unable to return to work and 40% have residual deficits (61). The present study will prospectively evaluate 200 LHC volunteers with a nested RCT to assess the impact of ivabradine on heart rate and symptoms in those meeting diagnostic criteria for two important types of autonomic dysfunction, POTS and inappropriate sinus tachycardia (IST). Our approach will include an integrated clinical, psychological and laboratory evaluation of a cohort of patients enriched for symptoms and signs consistent with autonomic dysfunction. A comparator group of 50 volunteers with documented COVID-19 but without LHC will be enrolled to better identify the risk factors for LHC and autonomic dysfunction. The comparator group will also be evaluated for POTS/IST and eligible for inclusion in the nested RCT if present.

A large number of interventions have been proposed for POTS. Standard of care remains limited and currently includes remedies such as compression garments, hydration and fludrocortisone to treat hypovolemia, drugs like midodrine to treat hypotension, and propranolol for tachycardia and increased sympathetic tone. Among other proposed experimental interventions such as intravenous immune globulin as a general anti-inflammatory and SSRIs for affective components, ivabradine has been proposed to lower heart rate and alleviate associated symptoms. Ivabradine is a unique specific modulator of sinus node automaticity that acts through specific blockade of the I_funny_ channel, whose expression is relatively restricted to cardiac nodal tissue. Other commonly used rate lowering agents including beta blockers and non dihydropyridine calcium channel blockers have off-target effects on cardiac contractility, blood pressure and/or bronchial tone. Limited evidence to date indicates that ivabradine may be effective for both POTS and IST by reducing inappropriately elevated heart rate in these conditions. Nevertheless, most of the data regarding ivabradine in POTS is anecdotal, with one small RCT demonstrating benefit to date (50). This study will add to the understanding of the potential therapeutic value of ivabradine in post-viral dysautonomia. Ivabradine’s current scope remains limited to the management of tachycardia associated with stable angina and heart failure not fully managed by beta blockers.

In addition to the primary aims, the study will contribute to identifying risk factors and better understanding pathophysiological processes, laying the groundwork for further exploration. The pathophysiologic basis and underlying mechanisms of disease remain poorly defined at present, though recent progress has been made (44). In the present study, we aim to take a multidisciplinary approach to better understand the diagnosis and management of LHC-associated POTS. LHC/POTS is believed to be caused by multiple pathophysiological processes including immune dysregulation, hematologic dysfunction, and dysautonomia. In our study, the risk factor evaluation will assess for hematological, immunological, and metabolic anomalies underlying the development of autonomic dysregulation. Genomic and multi-omic analysis will probe for genotypic and phenotypic risk factors for dysautonomia. Furthermore, symptoms of POTS/IST remain broadly heterogeneous, including overlapping psychological and psychiatric manifestations, for e.g., insomnia, depression, fatigue, memory loss. The study will incorporate evaluation of psychosocial, quality of life, and cognitive factors to better understand psychological and cognitive symptoms, and determine their interplay with physiologic responses to LHC.

Study of a complex disease with an intricate follow-up plan is bound to face practical implementation challenges. First, there is no established consensus for LHC diagnosis. Non specific symptoms such as fatigue might not prompt patients to seek medical care. As a result, LHC remains an underdiagnosed condition, thus, reducing the pool of potential patients available for recruitment. We aim to overcome this challenge through use of a comprehensive diagnostic strategy, to include the use of ambulatory measurements. Our recruitment strategy aims to combine community outreach including local healthcare providers to effectively identify and refer potential participants. Compliance with a host of study procedures and visits is a generic challenge for clinical research but may be compounded by disability and other challenges faced by POTS sufferers. Appropriate guidance and participant education will be key in ensuring successful adherence. The diagnostic complexity of POTS and related dysautonomia syndromes is also a potential limitation, as we have opted for an implementable subset of diagnostic tests from among the wide array available. Our hope is that use of multiple modalities to include ambulatory monitoring will ensure an adequate number of volunteers are enrolled in the RCT.

We estimate 20-25% of those with POTS-like symptoms presenting for the study will actually have POTS, but these could prove to be substantial over or under-estimates. Blinding a study using a rate-lowering agent may be challenging as well, particularly where placebo-treated subjects experience no benefit and are at risk for non-compliance or drop-out.

The management of LHC-associated POTS, a complex multifactorial condition, remains an emerging area of study and an ongoing challenge for patients and clinicians alike. The present study aims to add to the rapidly expanding LHC knowledge base, and explore one of several proposed interventions for LHC-associated cardiovascular autonomic dysfunction. We aim to accurately assess the effectiveness of ivabradine and its proper use in the management of LHC complicated by POTS. In so doing, we hope to provide a more solid evidence base for those struggling with this debilitating condition.

## Data Availability

All data produced in the present work are contained in the manuscript

## Trial status

This study is conducted under protocol IRB # USUHS.2022-094 approved in April 2022. The recruitment began on May 5^th^ 2023 and is expected to be completed within a year in May 2024.

**List of abbreviations**
ACE: Angiotensin Converting Enzyme
ANS: Autonomic Nervous System
CBC: Cell Blood Count
CGRP: Calcitonin Gene-Related Peptide
CIs: Confidence Intervals
CTCAE: Common Terminology Criteria for Adverse Events
COMPASS: Composite Autonomic Symptom Score
COVID-19: Coronavirus Disease 2019
CRFs: Case Report Forms
CRP: C - Reactive Protein
DASS-21: Depression Anxiety Stress Scale-21
DSMB: Data Safety Monitoring Board
DNA: Deoxyribonucleic Acid
DSM-V: Diagnostic and Statistical Manual of the DSM, 5th edition
EKG: Electrocardiogram
GAD: Generalized Anxiety Disorder
GPF: Gamma Prime Fibrinogen
FDA: Food and Drug Administration
HCN: Hyperpolarization-Activated Cyclic Nucleotide-Gated
HUTT: Head-Up Tilt-Table
IST: Inappropriate Sinus Tachycardia
LHC: Long-Haul’ COVID-19
MSAS: Memorial Symptom Assessment Scale
MHS: Military Health System
NIH: National Institutes of Health
NK: Natural Killer
ORs: Odds Ratios
PBMCs: Peripheral Blood Mononuclear Cells
PHQ: Patient Health Questionnaire
POTS: Postural Orthostatic Tachycardia Syndrome
PT: Prothrombin Time
PTT: Partial Thromboplastin Time
PROMIS: Patient-Reported Outcomes Measurement Information System
RCT: Randomized Controlled Trial
REDCap: Research Electronic Data Capture system
SA: Sinoatrial
SCID-5: Structured Clinical Interview for DSM-5 Disorders
SF-36: Short Form Health Survey
SNPs: Single Nucleotide Polymorphisms
TFTs: Thyroid Functions Tests
TMPRSS2: Transmembrane Protease Serine-2
VOSS: Vanderbilt Orthostatics Symptom Score
VWF: Von Willebrand Factor

## Declarations

### Ethics approval and consent to participate

The study was approved by the Uniformed Services University of the Health Sciences Institutional Review Board in June 2022 and registered with clinicaltrials.gov as NCT05481177.

Written informed consent will be obtained from all participants. A copy of the ethical approval and consent have been submitted as an additional file.

## Consent for publication

Not applicable.

## Competing Interest

The authors have no conflicts of interest to disclose.

## Funding

This study is funded by the Defense Health Agency (DHA), Department of Defense through the Uniformed Services University. Funds to perform the study were awarded through Cooperative Agreements with The Metis Foundation (Award # HU00012120090) and Henry M. Jackson Foundation for the Advancement of Military Medicine (Award # HU00012120087). The Funder has no role in the in study design; collection, management, analysis, and interpretation of data; writing of the report; or the decision to submit the report for publication.

## Author’s contribution

DS is the Principal Investigator and MH the Protocol Chair and grant awardee; they conceived the study, led the proposal and protocol development and drafted the manuscript. TA, JL, DB, KA, LH, RM, BM, DS, KP, PK, ME, AM, MTK, EG, EM, KP and CD assisted in the design of the development of the protocol as well as editing of the manuscript. DS, MH, BM, RM and STL drafted and edited the manuscript. MH supervised the design and implementation of the study and edited the manuscript. All authors read and approved the final manuscript.

## Acknowledgements

The authors would like to acknowledge our colleagues without whom the study would not have been possible. From the Uniformed Services University Drs. Jacob Collen, Marian Tanofsky Kraft, Ian Stewart, Paige Waterman, Martin Evans, Benjamin Pierson, Milissa Jones, Monique Hollis-Perry, Agnes Montgomery, Vivian Bauman, Katherine Thompson; Ms. Austin Pagani, Annette Cunningham, and Debra Weed. From the US Army Medical Research Institute of Infectious Diseases – Drs. Phillip Pittman, Erin Tompkins, Fernando Guerena and Ms. Carolyn Holland. From the Henry M. Jackson Foundation for the Advancement of Military Medicine Ms. Priscilla Kobi, Laurie Coran, Raquel Martinez, Heidi Adams, and Tabitha Shipmon. From the Metis Foundation – Marlene Barrera, Stephanie Margaret Mayne, Timothy Suzich, Joya Libbus, Keiko Fox, Holly Spinner, Isabel Thorstad, Hannah Repke, and Ashlee Simmons. From the Walter Reed Army Institute of Research Dr. Daniel Selig. From the Armed Forces Research Institute of Medical Sciences Drs. Pattaraporn Vanachayangul and Jeffrey Livezey. From the Walter Reed National Military Medical Center Drs. Jessica Wong-Flores, Nikhil Hauprikar and Wesley Campbell, and Mr. Victor Buckwold. From the US Army Research Institute of Environmental Medicine Dr. Geoff Chin and from the Joint Program Executive Office of Chemical and Biological Defense Dr. Anthony Cardile.

## Notes

### Competing Interest Statement

The authors have declared no competing interest.

### Clinical Trial

NCT05481177

### Funding Statement

This study is funded by the Defense Health Agency to the Uniformed Services University (Cooperative Agreement with The Metis Foundation) Award # HU00012120090".

